# Violence during pregnancy and mental health outcomes among women experiencing violence in rural Bangladesh

**DOI:** 10.1101/2025.08.02.25332863

**Authors:** Rifa Tamanna Mumu, Md Parvez Shaikh, Shadman Sakib Ayan

## Abstract

Violence during pregnancy is a significant public health issue, associated with serious mental health consequences, including antenatal depression and suicidal ideation. Although several studies in Bangladesh examined violence against women, there is a lack of research addressing its prevalence among pregnant women and the associated psychological outcomes.

This study aims to assess the prevalence of violence among pregnant women in rural Bangladesh and the prevalence of depression and suicidal ideation among affected individuals.

This study is a secondary analysis of cross-sectional data collected from January 8 to January 30, 2024, in a rural subdistrict in southern Bangladesh, involving 354 pregnant women. Data were collected using the Edinburgh Postnatal Depression Scale (EPDS) and analyzed with STATA version 17.

The prevalence of domestic violence (DV), lifetime intimate partner violence (IPV), and IPV during pregnancy were 5.9% (n = 21; 95% CI: 3.7%-8.9%), 5.4% (n = 19; 95% CI: 3.3%-8.3%), and 9.9% (n = 35; 95% CI: 7.0%-13.5%), respectively. Among women exposed to DV, 66.7% (n = 14; 95% CI: 42.6%-84.3%) reported antenatal depression. Similarly, depression was reported by 68.4% (n = 13; 95% CI: 42.7%-86.3%) of those with a history of lifetime IPV and 42.9% (n = 15; 95% CI: 27.1%-60.3%) of those experiencing IPV during pregnancy.

Suicidal ideation was present in 14.3% (n = 3; 95% CI: 4.2%-38.7%) of DV victims, 5.3% (n = 1; 95% CI: 0.6%-33.8%) of lifetime IPV victims, and 5.7% (n = 2; 95% CI: 1.3%-21.4%) of those experiencing IPV during their current pregnancy.

These findings reveal a concerning prevalence of violence against pregnant women and associated mental health issues. Urgent targeted interventions-including policy reforms, health education, and community-based awareness initiatives- are essential to address this critical public health challenge.

## Introduction

Violence against women is a major public health concern. One in three women worldwide experience physical abuse, rape, or other forms of violence in their lifetime (Ellsberg, 2006). The World Health Organization defines violence against women as any act perpetrated by a partner, family member, or community that causes physical, sexual, or psychological harm (World Health Organization, 2021). This includes a range of harmful practices such as sex-selective abortion, female genital mutilation, acid attacks, dowry-related deaths, honor killings (murdering women who is alleged shameful for the family), rape, and domestic violence (Watts & Zimmerman, 2002).

Domestic violence (DV) refers to psychological, physical, sexual, or economic abuse occurring within the family or between current or former partners, regardless of cohabitation status (Meyersfeld, 2012). Intimate partner violence (IPV), a subset of DV, encompasses physical, sexual, and emotional abuse, as well as controlling behaviors such as isolation and restriction. Forms of IPV include slapping, kicking, forced sex, humiliation, intimidation, and threats (World Health Organization, 2012).

The prevalence of DV across Asia ranges from 6.1% to 67.4% (Koirala & Chuemchit, 2020). In South Asia, it is highly prevalent: 40% of married Indian women and 88.8% married Pakistani women report experiencing some form of domestic violence (Hussain, Hussain, Zahra, & Hussain, 2020; Yoshikawa, Agrawal, Poudel, & Jimba, 2012). In Bangladesh, a study conducted during the COVID-19 pandemic found emotional violence at 19.9%, physical violence at 6.5%, and sexual violence at 3.0% (Hamadani et al., 2020).

Globally, 27.0% ever-partnered women aged 15-49 have experienced lifetime physical and/or sexual IPV (Sardinha, Maheu-Giroux, Stöckl, Meyer, & García-Moreno, 2022), with Southern Asia reporting the second-highest prevalence at 35.0% (World Health Organization, 2021). Bangladesh has the highest reported rate in the region, with 54.2% of women affected (Bhardwaj & Miller, 2021).

In Bangladesh, 42.4% of mothers with children under five have experienced IPV, which is associated with an increased risk of acute respiratory infections (ARI) and diarrhea in their children (Jay G. Silverman et al., 2009). IPV is particularly prevalent among pregnant women aged 15-35, leading to adverse outcomes such as low birth weight (LBW) and preterm births (Devries et al., 2010). Violence during pregnancy is not only common but also has farreaching physical and psychological consequences.

The effects of violence against women are profound. Research shows that DV significantly increases the risks of postpartum depression and postpartum blues among Asian women (Koirala & Chuemchit, 2020). Exposure to IPV is also a major risk factor for suicidal behavior, which remains a leading cause of female morbidity and mortality globally (Shoib et al., 2022). In Bangladesh, 13.8% of maternal deaths occur due to violence, and nearly 25.0% of women experience pregnancy loss—in the form of miscarriage, induced abortion, or stillbirth—linked to IPV (Hossain, 2016; Jay G Silverman et al., 2007).

DV and IPV also impact family dynamics and child development. Children of DV victims often experience behavioral and mental health problems, while families bear both psychological and economic burdens (A. R. J. A. S. W. Khan & Review, 2015).

Although several studies have examined DV and IPV in Bangladesh, limited research exists on the prevalence among pregnant women and the associated risk of depression and suicidal ideation. This study aims to address that gap by estimating the prevalence of DV, lifetime IPV, and IPV during pregnancy, as well as the mental health consequences—specifically depression and suicidal ideation—among affected pregnant women in rural southern Bangladesh.

A clear understanding of these issues can guide the development of strategic policies, community interventions, and mental health services to combat violence, safeguard women’s rights and dignity, and promote psychological well-being.

## Materials and methods

### Study design and setting

This study is a secondary analysis of cross-sectional data collected between January 8 to January 30, 2024, at two healthcare facilities in Lohagara, a rural sub-district of Narail in southern Bangladesh. The facilities included the Upazila Health Complex (a government hospital) and Khan General Hospital, Lahuria (a private hospital).

### Study participants, Sample size, and sampling technique

The original study included pregnant women at any trimester attending antenatal checkups at both hospitals during the study period. Participants were selected through systematic sampling. Every third patient attending antenatal care in both hospitals was invited for a face-to-face interview. The sample size was calculated using the formula: 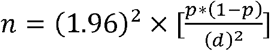.

A total of 354 participants completed the survey.

### Data collection, measurement tools, management & analysis

Data were collected using two structured questionnaires, translated into Bengali. The first included sociodemographic and health-related questions. The second was the validated Bengali version of the Edinburgh Postnatal Depression Scale (EPDS-B) [22-24] to evaluate antenatal depression. The EPDS has 10 items with a total score ranging from 0 to 30. A score of 10 or higher indicated probable antenatal depression [25-27]. Suicidal ideation was assessed based on a score above 0 on the final EPDS item.

Patients who were non-resident, had cognitive difficulties, and were diagnosed with psychiatric disorders were excluded. A 15-minute interview was conducted in a separate room to ensure confidentiality. Eligible participants provided written informed consent before the interview. Data were reviewed daily for accuracy and completeness; errors were corrected, and missing data were handled by listwise deletion. We used STATA version 17 for analysis.

## Result

### Sociodemographic characteristics of participants

A total of 354 data points were collected. The majority of participants (92.9%, n = 328) were between 18 and 35 years of age. Over half (53.8%, n = 190) were in their second trimester of pregnancy. A small proportion (7.3%, n = 26) had no formal or primary-level education, and most were unemployed (96.1%, n = 340). Approximately one-quarter (26.3%, n = 93) reported a monthly household income below 10,000 BDT. The vast majority of participants (96.3%, n = 341) identified as Muslim, and nearly half (48.6%, n = 172) had married before the age of 18. Error! Reference source not found..

### Prevalence of violence against pregnant women in Bangladesh

As shown in Error! Reference source not found., the prevalence of domestic violence (DV) among pregnant women was 5.9% (n = 21; 95% CI: 3.7%-8.9%). The lifetime prevalence of intimate partner violence (IPV) was 5.4% (n = 19; 95% CI: 3.3%-8.3%), while IPV during the current pregnancy was 9.9% (n = 35; 95% CI: 7.0%-13.5%).

### Prevalence of antenatal depression among pregnant women experiencing violence

Error! Reference source not found. Illustrates that antenatal depression was reported by 66.7% (n = 14; 95% CI: 42.6%-84.3%) of those who experienced DV, 68.4% (n = 13; 95% CI: 42.7%-86.3%) of those with a history of lifetime IPV, and 42.9% (n = 15; 95% CI: 27.1%-60.3%) of those who experienced IPV during pregnancy. Additional information on the underlying data is provided in the supplementary materials.

### Prevalence of suicidal ideation among pregnant women experiencing violence

As presented in Error! Reference source not found., the prevalence of suicidal ideation among DV victims was 14.3% (n = 3; 95% CI: 4.2%-38.7%). Among those reporting lifetime IPV, the prevalence was 5.3% (n = 1; 95% CI: 0.6%-33.8%), and among those experiencing IPV during pregnancy, it was 5.7% (n = 2; 95% CI: 1.3%-21.4%).

## Discussion

This study assessed the prevalence of violence against pregnant women and examined antenatal depression and suicidal ideation among victims in rural southern Bangladesh. The findings indicate that 5.9% (n = 21; 95% CI: 3.7%-8.9%) of participants experience domestic violence (DV), 5.4% (n = 19, 95% CI: 3.3%-8.3%) report lifetime intimate partner voilence (IPV), and 9.9% (n = 35, 95% CI: 7.0%-13.5%) experience IPV during pregnancy. Among women exposed to DV, 66.7% (n = 14; 95% CI: 42.6%-84.3%) report antenatal depression. The prevalence of depression is 68.4% (n = 13; 95% CI: 42.7%-86.3%) among those with a history of lifetime IPV and 42.9% (n = 15; 95% CI: 27.1%-60.3%) among those exposed to IPV during pregnancy. Suicidal ideation is 14.3% (n = 3; 95% CI: 4.2%-38.7%) among DV victims, 5.3% (n = 1; 95% CI: 0.6%-33.8%) among those with lifetime IPV, and 5.7% (n = 2; 95% CI: 1.3%-21.4%) among those currently experiencing IPV.

In South Asia, women are subjected to various forms of violence, including physical assault, sexual violence, abduction for prostitution, and honor killings. Factors contributing to this violence include dowry practices, male-dominated family structures, patriarchal norms, and rigid cultural traditions (Niaz, 2003). In Pakistan, 51.0% of women experience DV in the six months before or during pregnancy, and 16.0% develop suicidal ideation as a result (Karmaliani et al., 2008). In Bangladesh, the COVID-19 pandemic has exacerbated these issues, with DV prevalence rising to 36.8% (Rashid Soron et al., 2021). Contributing factors include low socioeconomic status, large family size, indebtedness, food insecurity, and exposure to crisis events (Haque et al., 2022).

In parts of India and Pakistan, women are often treated as second-class citizens, and minor mistakes are met with severe punishment (Niaz, 2003). In India, the prevalence of physical abuse is estimated at 34.0%, with 15.0% experiencing violence during pregnancy. Among abused women, 72.0% report depression (Fikree, Bhatti, & Obstetrics, 1999). During the COVID-19 pandemic, approximately 45.0% of Bangladeshi women reported experiencing IPV (Rayhan & Akter, 2021).

The prevalence of IPV during pregnancy in South Asian Countries is 23.4%(Nepal et al., 2022). In a study, Jain et al. identify a 12.3% prevalence of IPV during 20-28 weeks of pregnancy in India, where male child preference is a significant contributing factor (Jain et al., 2017). In Karachi, Pakistan, 44.0% of women experience spousal abuse during pregnancy (Farid, Saleem, Karim, & Hatcher, 2008). A pre-pandemic study in two eastern subdistricts of Bangladesh has identified a 66.4% prevalence of IPV during the first six months of pregnancy. Risk factors include dowry demands, lack of social support, husbands’ controlling behavior, low self-esteem, limited decision-making autonomy, and childhood exposure to violence (M. J. Islam, Mazerolle, Broidy, & Baird, 2021).

Approximately 90.0% of Bangladesh’s population is Muslim [36]. Although Islam prohibits cruelty and family violence, patriarchal interpretations often misrepresent religious teachings. In some rural communities, religious leaders enforce social norms under the guise of religion, sometimes justifying abuse [37, 38]. Many rural families lack awareness of the physical and mental health needs of pregnant women. Some families tolerate or even justify violence against expectant mothers in response to perceived misconduct or the likelihood of giving birth to a female child.

On the other hand, a significant portion of Bangladeshi women internalize these norms: nearly one-third believe that being beaten by a husband is justified (M. N. Khan & Islam, 2018). Women exposed to violence in childhood are more likely to tolerate IPV (Vung, Ostergren, & Krantz, 2008). One study found that 20.5% of Bangladeshi women approve of at least one form of IPV, a view strongly associated with low education and conservative religious beliefs (M. Islam, Ahmed, & Mistry, 2021). Factors such as fear of retaliation, concern for children, lack of financial or social support, emotional attachment, and hope for husband’s behavioral change often compel women to remain with abusive partners (World Health Organization, 2012). Other contributing factors include dowry disputes, substance abuse by husbands, sexual coercion, infertility, the birth of female children, and entrenched patriarchal values (Hossain, 2016).

IPV is associated with poor maternal mental health. DV is associated with antenatal and postnatal anxiety, depression, and post-traumatic stress disorder (PTSD) (Howard, Oram, Galley, Trevillion, & Feder, 2013). A study in India highlights that childhood physical or sexual abuse is associated with higher rates of antepartum depression (Barrios et al., 2015). IPV during pregnancy is significantly associated with prenatal depression and adverse birth outcomes among the Chinese population (Yu et al., 2018). In rural Bangladesh, antenatal depression affects approximately 39.0% of pregnant women, with IPV identified as a key risk factor.

Suicide is a leading cause of maternal death globally (Chin, Wendt, Bennett, & Bhat, 2022). One study suggests that 20% of postpartum deaths occur due to suicide (Lindahl, Pearson, & Colpe, 2005). In Bangladesh, 11.0% of pregnant women experience suicidal thoughts, and 6.5% attempt suicide (Li, Imam, Jing, Wang, & Zhou, 2020). Exposure to IPV during pregnancy significantly increases the risk of suicidal ideation (Gelaye, Kajeepeta, & Williams, 2016).

This secondary analysis uses cross-sectional data collected from two hospitals at a remote southern subdistrict of Bangladesh. Data were collected from both government and non-government hospitals using a systematic sampling technique to capture the diversity of the population. Participants were selected regardless of their background, education, employment, and socio-economic status. However, in the context of rural Bangladesh, there might be a subset of the population who do not attend antenatal care unless they face severe physical challenges. For this reason, this study may be subject to a generalizability bias.

Additionally, as this is a secondary analysis, we have limited control over modifying or expanding the previous data collection process to address the current research objectives.

## Conclusion

The prevalence of violence among pregnant women in rural Bangladesh remains a serious public health concern, with profound implications for maternal mental health. Approximately one in twenty pregnant women experiences domestic violence, and nearly one in ten report intimate partner violence (IPV) during pregnancy. Among those exposed to violence, at least 40.0% suffer from antenatal depression, and over 5.0% report suicidal ideation.

Despite the gravity of the issue, limited measures have been implemented to address it. To reduce the burden of antenatal depression and suicidal thoughts, it is imperative to prevent violence against women. This requires the enforcement of robust legislation that protects women’s rights and holds perpetrators accountable. Empowering women through education is equally critical, as it fosters self-confidence, awareness of legal and social rights, and a sense of dignity.

While the government has made efforts to promote women’s empowerment, these initiatives must be expanded to the grassroots level. Policies targeting the reduction of child marriage and school dropout rates should be prioritized. Community-based health education programs and awareness campaigns must be implemented to engage women, families, and communities in addressing the issue.

A coordinated, multisectoral response is essential. Policymakers, law enforcement agencies, healthcare professionals, social activists, and local leaders should work collaboratively to develop targeted policies, implement educational initiatives, and increase public awareness. Hospitals should integrate mental health support into antenatal care, and community health workers should play a proactive role in educating families about the importance of regular antenatal visits and mental well-being during pregnancy.

Through sustained, collaborative efforts, it is possible to significantly reduce violence against pregnant women and mitigate its devastating effects on mental health.

## Data Availability

Data is available on figshare.com.
Characteristics of pregnant women in Lohagara, Narail, 2024. DOI: 10.6084/m9.figshare.24994110v3

https://figshare.com/articles/dataset/_b_Prevalence_and_associated_factors_of_antenatal_depression_in_rural_Bangladesh_b_/24994110?file=56151293

---

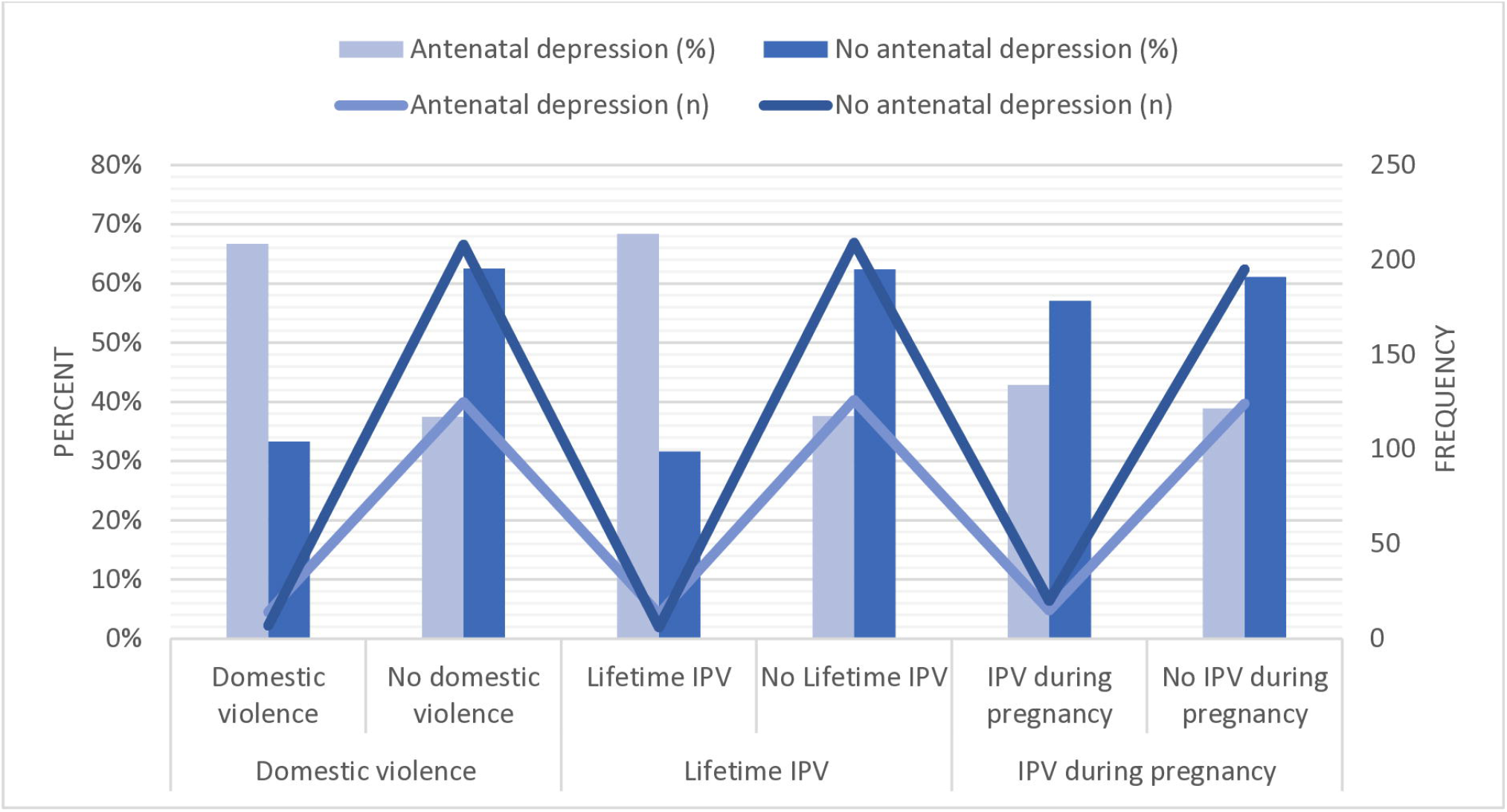

