## Supplementary material for "Violence during pregnancy and mental health outcomes among women experiencing violence in rural Bangladesh": Table 1

**Table 1 Baseline characteristics of pregnant women (n = 354)**

| Variables | Category | Frequency (n) | Percent (%) |
| --- | --- | --- | --- |
| Age | < 18 years | 20 | 5.7 |
|  | 18–35 years | 328 | 92.9 |
|  | > 35 years | 5 | 1.4 |
| Gestational weeks | < 12 weeks | 56 | 15.9 |
|  | 13–28 weeks | 190 | 53.8 |
|  | > 13 weeks | 107 | 30.3 |
| Education | Educated | 328 | 92.7 |
|  | Not educated | 26 | 7.3 |
| Employment | Employed | 14 | 3.9 |
|  | Unemployed | 340 | 96.1 |
| Monthly family income (BDT) | < 10000 | 93 | 26.3 |
|  | 10,000–20,000 | 205 | 57.9 |
|  | > 20,000 | 56 | 15.8 |
| Religion | Hindu | 13 | 3.7 |
|  | Muslim | 341 | 96.3 |
|  | Others | 0 | 0.0 |
| Marital age | < 18 years | 172 | 48.6 |
|  | 18 years or more | 182 | 51.4 |
