## Supplementary material for "Violence during pregnancy and mental health outcomes among women experiencing violence in rural Bangladesh": Table 2

**Table 2 Prevalence of violence against pregnant women (n = 354)**

| Types of violence | Category | Number (n) | Percentage (%) | 95% CI (%) |
| --- | --- | --- | --- | --- |
| Domestic violence | Yes | 21 | 5.9 | 3.7–8.9 |
|  | No | 333 | 94.1 | 91.1–96.3 |
| Lifetime IPV | Yes | 19 | 5.4 | 3.3–8.3 |
|  | No | 335 | 94.6 | 91.7–96.7 |
| IPV during pregnancy | Yes | 35 | 9.9 | 7.0–13.5 |
|  | No | 319 | 90.1 | 86.5–93.0 |
