## Supplementary material for "Violence during pregnancy and mental health outcomes among women experiencing violence in rural Bangladesh": Table 3

**Table 3 Suicidal ideation among pregnant women experiencing violence (n=354)**

| Types of violence | Category | Suicidal ideation | | | | | |
| --- | --- | --- | --- | --- | --- | --- | --- |
|  |  | Yes | | | No | | |
|  |  | Frequency  (n) | Percent  (%) | 95% CI | Frequency  (n) | Percent  (%) | 95% CI |
| Domestic violence | Yes | 3 | 14.3 | 4.2%–38.7% | 18 | 85.7 | 61.3%–95.8% |
|  | No | 40 | 12.0 | 8.9%–16.0% | 293 | 88.0 | 84.0%–91.1% |
| Lifetime IPV | Yes | 1 | 5.3 | 0.6%–33.8% | 18 | 94.7 | 66.2%–99.4% |
|  | No | 42 | 12.5 | 9.4%–16.6% | 293 | 87.5 | 83.4%–90.6% |
| IPV during pregnancy | Yes | 2 | 5.7 | 1.3%–21.4% | 33 | 94.3 | 78.6%–98.7\|% |
|  | No | 42 | 12.9 | 9.6%–17.0% | 278 | 87.2 | 83.0%–90.4% |
